# Sex-specific transcriptional differences and loss of gene imprinting in pancreatic neuroendocrine tumors

**DOI:** 10.1101/2021.06.09.21258573

**Authors:** Nikolay A. Ivanov, Kirill Grigorev, Thomas J. Fahey, Brendan M. Finnerty, Christopher E. Mason, Irene M. Min

**Affiliations:** Department of Surgery, Weill Cornell Medicine, New York, New York, USA; Weill Cornell Medical College, New York, New York, USA; Clinical & Translational Science Center, Weill Cornell Medicine, New York, New York, USA; Department of Physiology and Biophysics, Weill Cornell Medicine, NY, USA; The HRH Prince Alwaleed Bin Talal Bin Abdulaziz Alsaud Institute for Computational Biomedicine, Weill Cornell Medicine, NY, USA

## Abstract

Pancreatic neuroendocrine tumors (PNETs) occur more frequently in men and are associated with higher mortality in males; however, the molecular basis for these sexual dimorphisms is unclear. Here, we demonstrate that PNETs are associated with the emergence of unique sex-specific transcriptomic differences that are not observed in non-neoplastic pancreatic islet tissues. We also show that while widespread sex-specific differences are present in the DNA methylation landscapes of control pancreatic islets, they are erased in PNETs. This includes a loss of imprinting with regards to many genes. These results implicate an emergence of sex-associated genetic and epigenetic dysregulations in PNETs.

---

Pancreatic neuroendocrine tumors (PNETs) are rare neoplasms that develop from pancreatic neuroendocrine cells located in the islets of Langerhans. They represent the second most common primary pancreatic neoplasms and make up approximately 2% of all pancreatic tumors (1). Epidemiologic studies demonstrate that PNETs exhibit sexual dimorphisms with regards to prognosis, disease recurrence, and complication rates (2). Male predominance in PNET incidence has been observed in the Surveillance, Epidemiology, and End Results (SEER) database for decades (3, 4). Men also exhibit lower overall survival (5), higher cancer-specific mortality (6), and significantly lower 5-year survival probability than women (7). For instance, an analysis of 7,946 cases demonstrated that 55.6% of cases occur in men (3), an examination of 131 cases showed that 5-year survival probability was 2.8-fold higher in females (p=0.02) (8), and a study of 653 cases in elderly patients (75 years old) showed that female patients had a 28% reduction in risk of death due to cancer (HR = 0.72 in females; 95% CI: 0.56–0.93) (6).

While it has been shown that estrogen and progesterone affect PNET development (9-12), genome-wide sex-specific differences in PNETs have not been explored. We therefore sought to investigate the sexual dimorphisms in transcription and DNA methylation landscapes of PNETs to identify novel genomic sex variation that may be associated with the observed clinical differences.

We generated RNA sequencing data from 21 primary PNETs (9 Female (F), 12 Male (M)) (Supplementary Table 1) surgically resected at our institution (Discovery Dataset). To validate our results, we downloaded a publicly available PNET RNAseq data from the Sequence Read Archive (SRA), consisting of 24 primary PNETs (10 F, 14 M) (Validation Dataset) (Supplementary Table 1). To compare PNET data to controls, we further downloaded RNAseq data from 89 non-neoplastic (control) pancreatic islet tissue samples (35 F, 54 M) (Supplementary Table 2).

In our discovery dataset, we found 559 genes differentially expressed (DE) by sex at genome-wide significance (FDR ≤ 0.1), with 60.5% of them overexpressed in males. In the validation dataset, 659 genes were DE by sex, with 60.1% overexpressed in males. In the control dataset, only 173 genes were DE by sex (60.1% overexpressed in males) (Figure 1a-c). Interestingly, in both the discovery and validation PNET datasets, there were significantly more genes differentially expressed by sex as compared to control pancreatic islet tissues (discovery vs control: p = 6.5×10^−5^; validation vs control: p = 2.0×10^−48^), despite greater sample size (and therefore power) in the control dataset (Figure 1a and Supplementary Table 3). Additionally, there were significantly more transcription factors (TFs) differentially expressed between sexes in the PNET datasets than in control data (55 and 65 differentially expressed TFs in the discovery and validation datasets respectively; 15 TFs in the control dataset) (discovery vs control: p = 2.7×10^−6^; validation vs control: p = 3.4×10^−8^). Eight TFs were differentially expressed by sex in both discovery and validation datasets, and while 5 of them are located on sex chromosomes and were also differentially expressed by sex in controls, 3 autosomal TFs (HSPA1A, SOX15, and APOA4) were uniquely DE by sex only in PNET samples (see Supplementary Table 3). Notably, SOX15 plays an important role in tumorigenesis in various cancers (13), potentially including pancreatic cancer (14).

**Figure 1.**
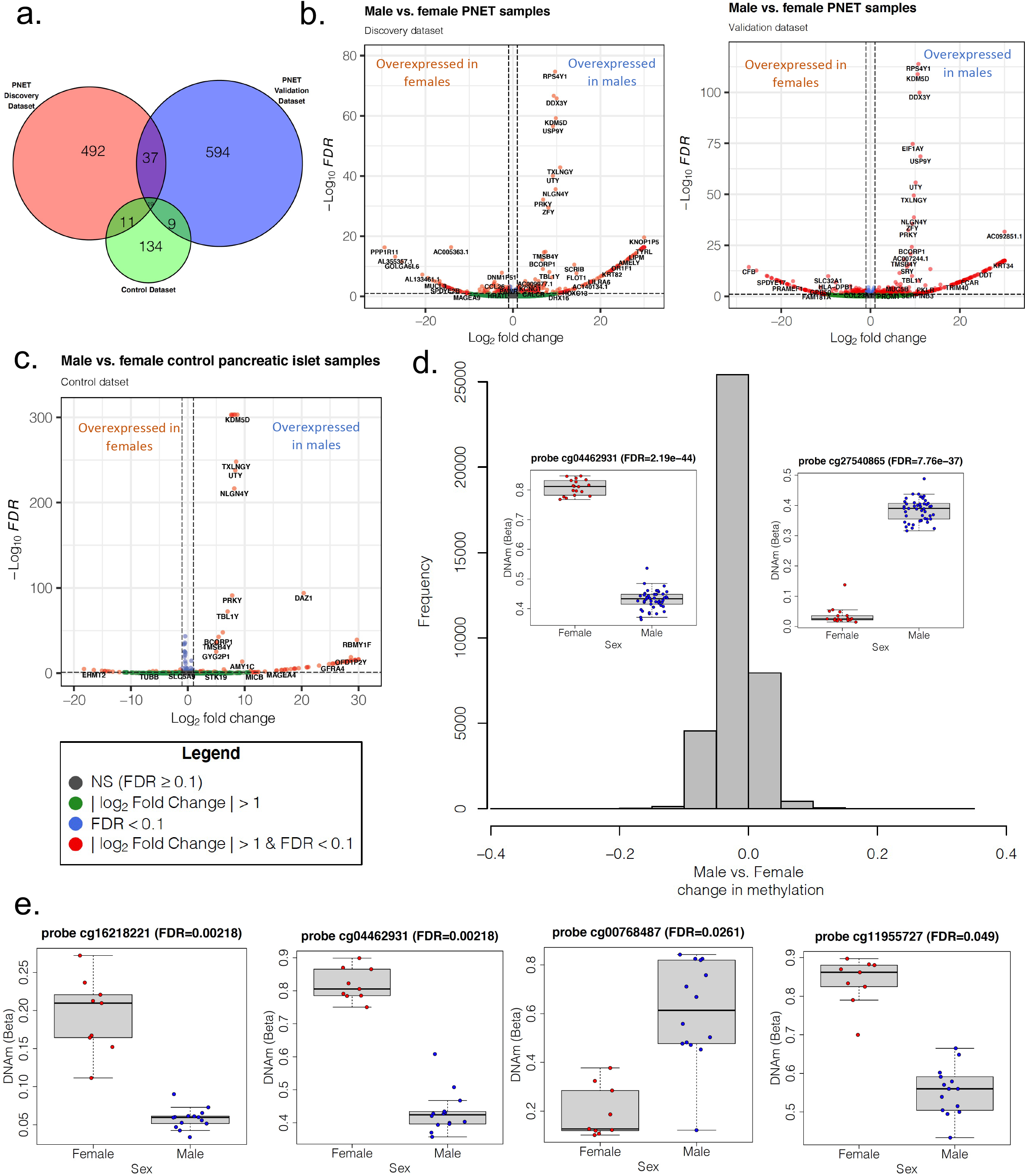
Sex-specific differences in PNET genomic profiles. **a**: Area-proportional Venn diagram showing overlap of genes differentially expressed by sex (at FDR 0.1) in the discovery and validation PNET datasets and control pancreatic islets. **b, c**: Volcano plots of genes differentially expressed by sex in PNETs (b) and control pancreatic islets (c). Genes with an FDR < 0.1 and an absolute effect size (|log_2_FC|) > 1 are indicated in red. **d**: Histogram of difference in DNAm levels at CpGs/probes differentially methylated between male- and female-derived control pancreatic islet tissue samples (at FDR 0.1). The x-axis shows the difference in DNAm level (on the proportion methylation (Beta) scale) between male- and female-derived samples. The insets show representative cytosines differentially methylated by sex. **e**: Cytosines differentially methylated by sex in PNETs. The y-axes denote DNAm levels on the proportion methylation (Beta) scale.

In order to investigate whether specific biological programs are differentially regulated by sex, we performed gene set enrichment analysis on the DE genes in our discovery dataset using goseq (see Methods). We found significant enrichment for the Gene Ontology (GO) biological process (BP) ‘detoxification of copper ion’ (FDR = 0.019). Interestingly, copper transport has been previously implicated in cancer development (15). In the validation dataset, there was significant enrichment for the GO BP ‘lipoprotein metabolic process’ (FDR = 0.0079), which has also been linked to oncogenesis (16).

Of all the genes differentially expressed by sex in PNETs, 56 were found to be differentially expressed in both the discovery and validation cohorts. Of these, 37 were differentially expressed by sex in PNETs but not in controls. (All 37 genes are located on autosomal chromosomes.) Sixteen of these genes exhibited concordant fold-changes between the discovery and validation cohorts, meaning that they were differentially expressed by sex in the same direction (i.e., overexpressed in the same sex) in both datasets (Table 1). Some of these genes play known roles in tumorigenesis. For instance, *RASSF7* (overexpressed in males with a large effect size: log_2_FC of 30 and 23 in the discovery and validation analyses respectively) is an oncogene that promotes non-small cell lung cancer development and metastasis (17). *RXRB* is another gene that we found to be overexpressed in males, and it has been shown to promote cancer stemness by serving as a major effector of the oncogene *RAB39A* in the *RAB39A—RXRB* axis (18). Not all differentially expressed genes favored worse outcomes in males. *CD52*, overexpressed in female PNETs, is a lymphocyte surface glycoprotein known to inhibit effector T-cells (19). It is upregulated in breast cancer and is associated with reduced survival rate (20). The mitogen *IGF2*, whose expression was significantly overexpressed in male-derived PNET tissues, is known to be upregulated in various tumors (21), and has been shown to drive the development of neuroendocrine tumors (22).

**Table 1.** Genes differentially expressed by sex in concordance* in both the discovery and validation PNET datasets.

|  | Discovery PNET dataset |  | Validation PNET dataset |  |
| --- | --- | --- | --- | --- |
| Gene | log2 Fold Change ** | FDR | log2 Fold Change ** | FDR |
| *SPDYE2B* | -15.52 | 4.09E-04 | -6.49 | 6.50E-02 |
| *CLVS2* | -5.26 | 8.23E-04 | -2.60 | 8.51E-03 |
| *CLEC10A* | -1.89 | 4.24E-03 | -0.55 | 3.55E-02 |
| *CD52* | -1.65 | 4.54E-02 | -0.70 | 1.73E-03 |
| *TMEM273* | -0.89 | 3.23E-02 | -0.52 | 7.92E-03 |
| *GRASP* | 1.89 | 3.06E-02 | 3.79 | 7.25E-02 |
| *TSHR* | 2.76 | 7.94E-02 | 6.06 | 9.18E-02 |
| *APOBEC2* | 3.24 | 1.11E-02 | 8.09 | 6.54E-02 |
| *IGF2* | 4.19 | 2.68E-03 | 10.01 | 1.53E-02 |
| *ANXA13* | 4.91 | 1.38E-03 | 7.02 | 5.87E-02 |
| *APOA4* | 5.41 | 2.47E-02 | 17.65 | 8.57E-02 |
| *AC132217.2* | 6.06 | 8.18E-02 | 12.50 | 6.46E-03 |
| *RXRΒ* | 9.60 | 3.69E-02 | 17.80 | 7.67E-02 |
| *AC136759.1* | 11.84 | 2.70E-02 | 22.96 | 2.25E-02 |
| *TBC1D3G* | 21.68 | 1.05E-08 | 30.00 | 2.40E-03 |
| *RASSF7* | 30.00 | 4.30E-17 | 22.65 | 1.86E-02 |

\* Concordance refers to genes being differentially expressed by sex in the same direction (i.e., overexpressed in the same sex) in both the discovery and validation datasets.
\*\* log2 Fold Change > 0 denotes gene overexpression in males

*IGF2* is a paternally-expressed imprinted gene (21). Because DNA methylation is closely associated with gene expression, we hypothesized that genes differentially expressed by sex may be regulated by DNA methylation. To investigate this hypothesis, we downloaded publicly available DNA methylation (DNAm) data (see Methods) obtained with the Illumina HumanMethylation450 microarray (Illumina 450k) (23).

We obtained matched DNAm – gene expression data for 23 primary PNET samples (9 F, 14 M) (Supplementary Table 1) and DNAm data for 64 non-neoplastic (control) pancreatic islet tissue samples (18 F, 46 M) (Supplementary Table 2). We analyzed data across 456,928 probes targeting single methylation loci after excluding 17,120 loci with common single nucleotide polymorphisms and probes on the sex chromosomes (see Methods).

We found widespread sex specific DNAm differences in the control pancreatic islet dataset, with 38,623 cytosines (8.4% of all interrogated probes) differentially methylated by sex at genome-wide significance (FDR 0.1) (Figure 1d). Of note, since the Illumina 450k array probes target single cytosines (CpGs or to a much lesser extent CHs), differentially methylated cytosines are also commonly referred to as a differentially methylated probes (DMPs). Many of the sex-DMPs (16,800) were associated with regulatory elements (promoters or enhancers) and 35,611 overlapped or were within 5 kb of genes (Supplementary Table 4). None of the sex-DMPs clustered together into differentially methylated regions (see Methods). There were also no long-range regions with consistent significant methylation changes between sexes (DNA methylation ‘blocks’) (see Methods).

In contrast to controls, we found sparse DNAm differences by sex in PNET samples, with only 4 cytosines differentially methylated by sex at genome-wide significance (Figure 1e), 3 of which were also differentially methylated by sex in controls. Of the 4 sex-specific sex-DMPs, 3 overlapped or were within 5 kb of at least one gene (Supplementary Table 5), however, none of these proximal genes exhibited significant correlation between DMP methylation levels and gene expression (Supplementary Figure 2).

As expected, *IGF2* was differentially methylated by sex in the control dataset (4 sex-DMPs overlapped *IGF2*, see Supplementary Table 5). However, *IGF2* was not differentially methylated in the PNET dataset, suggesting a loss of imprinting in the PNET state. In the control dataset, 605 sex-DMPs overlapped or were within 5kb of 87 unique imprinted genes (Supplementary Table 5), whereas no imprinted genes were differentially methylated by sex in PNET samples.

In conclusion, we show that sex-specific differences emerge in PNET transcriptomes. Gene expression in pancreatic islets is known to differ by sex (24), however, our analysis indicates that the transcriptional differences in PNETs are distinct from those that are observed in non-neoplastic tissue counterparts. Interestingly, these PNET gene expression differences by sex are not related to DNA methylation. In fact, while control islets harbor >38,600 sex-specific methylome differences, they appear to be almost completely erased in the cancer state.

Effects of sex on tumorigenesis and clinical outcomes has emerged as an important component in our understanding of oncologic disease (25, 26). However, as yet, there are no sex-specific cancer therapy protocols and no clinical trials investigating outcome differences by sex. It is therefore the logical next step to identify sex-driven differences that would affect prognosis and response to therapy and incorporate that information into clinical trial design. Thus, achieving a greater understanding of molecular differences in PNETs by sex is invaluable for advancing personalized therapy and informing future clinical trial design. Lastly, pancreatic neuroendocrine tumors encompass several heterogeneous subgroups, with distinct genomic, clinical, and prognostic profiles. Future studies should analyze sex differences in these PNET subtypes individually, yielding results with a high degree of internal validity, which are applicable to a narrow range of patients with specific cancer subtypes, a concept that underlies the essence of personalized medicine.

## Methods

### Human PNET tissue collection

Studies were conducted in accordance with the Declaration of Helsinki. Informed written consents were obtained from patients with a diagnosis of PNETs pre-operatively for study enrollment under the Weill Cornell Medicine Institutional Regulatory Board approval. Clinical specimens (derived from primary PNETs) were collected from patients undergoing surgery at Weill Cornell Medicine/NewYork Presbyterian Hospital.

### Tumor RNA sequencing data generation

Total RNA was extracted from flash-frozen tumor tissues using the materials and protocol provided in the RNeasy mini kit (Qiagen). RNA yield was measured using the NanoDrop Microvolume Spectrophotometer (ThermoFisher Scientific, Waltham, MA).

Total RNA was used for TruSeq stranded library generation, and paired end clustering with 51×2 cycles of sequencing performed on HiSeq 2500/4000 (Illumina) by the Genomics Core Facility of the Weill Cornell Medicine Core Laboratories Center.

### RNA sequencing data download

Raw PNET RNAseq validation data (FASTQ files) was downloaded from SRA (www.ncbi.nlm.nih.gov/sra; BioProject ID PRJNA484008), as was raw RNAseq data from control pancreatic islet tissue (BioProject: PRJNA217347). For PNET data, only samples generated from primary tumors were used; samples taken from metastatic sites were not utilized. Md5 checksums were verified for all downloaded FASTQ files to ensure download fidelity.

### RNA sequencing data processing

RNA sequencing reads (FASTQ files) were quantified using *Salmon* (27), correcting for fragment-level GC biases (28) and using an Ensembl human transcriptome (release 97). A pre-built full decoy transcriptome index for Human Genome version hg38 (downloaded from www.refgenomes.databio.org/) was utilized when running *Salmon*.

Quantified data was then imported into R and transcript-level quantification was summarized to gene-level with the *tximeta* R package (version 1.8.4). Differential expression analysis was performed with *DESeq2* (version 1.30.0) (29) and results were extracted via the DESeq2 *results* function using the default settings. To adjust for significant variation (due to factors other than the covariate of interest) surrogate variables were computed with the *SVA* R/Bioconductor package (version 3.38.0) and adjusted for (30). 6 significant Surrogate Variables (SVs) were detected in our discovery and validation PNET analyses and 14 SVs were detected in the control tissue analysis. Genes with Benjamini-Hochberg adjusted p-values (FDR) ≤ 0.1 were called significantly differentially expressed by sex. Gene annotations in all analyses were based on Ensembl release 97.

To determine which differentially expressed genes were human transcription factors, a comprehensive list of human transcription factors was obtained from a review article by Lambert et al (http://humantfs.ccbr.utoronto.ca/allTFs.php) (31).

*Salmon* quantification was performed on Linux (Red Hat Enterprise Linux Server release 6.3); all other analyses were done in R (version 4.0.3).

### Gene set over representation analysis

Over Representation Analysis (ORA) for Gene Ontology terms and KEGG pathways was performed with the *goseq* Bioconductor package (version 1.42.0) (32), adjusting for gene length. The probability weighing function (PWF) was calculated with the data bias (*bias*.*data* argument in the *nullp* function) set to the median transcript length for each gene. The median transcript length of each gene was calculated from Ensembl (release 97) data. Category enrichment scores were calculated using the Wallenius distribution to approximate the null distribution. FDR 0.1 was considered statistically significant. Analyses were done in R (version 4.0.3).

### DNA methylation data procurement

Raw Illumina HumanMethylation450 microarray data (IDAT files) was downloaded from the Gene Expression Omnibus (GEO). (PNET data: GEO accession number GSE117852; control pancreatic islet data: GEO accession number GSE143209). For PNET data, only samples generated from primary tumors were used; samples taken from metastatic sites were not utilized.

### DNA methylation data processing and normalization

Raw DNAm data (Red and Green channel intensities from the array probes) was converted into M(ethylated) and U(nmethylated) signals using the Bioconductor package *minfi* (33) (version 1.36.0). To perform quality control, we examined the log_2_ of median M and U intensities for all samples. Samples that have low median methylated and unmethylated intensities are typically flagged as likely low-quality samples (33); none of our samples had to be flagged for low quality (see Supplementary Figure 3).

Next, we converted the Red and Green intensities into a methylation signal without using any normalization (via minfi’s *preprocessRaw* function) and examined the distribution of proportion methylation (β) values for all samples (see Supplementary Figure 4). Of note, β-values are computed for the *i*^*th*^ interrogated cytosine via the following equation:

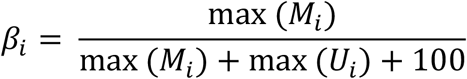

where M_i_ and U_i_ are intensities measured by the *i*^*th*^ Methylated and Unmethylated probes, respectively, and 100 is an offset constant which is added to regularize the β-value when M and U intensities are low, per Illumina’s recommendation (34). All of our samples exhibited a bimodal distribution of β-values, with enrichment of low β-values (close to zero) and high β-values (close to 1) (Supplementary Figure 4), consistent with expected results (33).

Next, the data was normalized via functional normalization, an unsupervised method which removes unwanted technical variation by utilizing control probes on the array. Functional normalization was shown to outperform existing normalization methods when analyzing data that is expected to have global DNAm differences, such as cancer data (35). After normalization, we removed 17,120 probes that contained common SNPs (based on dbSNP build 147) at target cytosines or single base extension (SBE) sites. We also dropped the probes on sex chromosomes, as they are harder to accurately normalize (36). At the end of data processing, we ended up with in a total of 456,928 probes (each one targeting a unique CpG/CH) on 87 samples.

### Differential methylation analysis

To find differentially methylated cytosines (referred to as Differentially Methylated Positions/Probes, or DMPs) we fit the following linear model for every cytosine interrogated by the array:

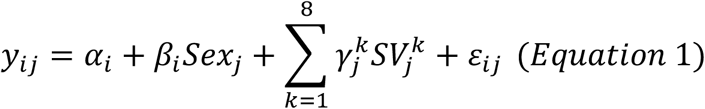

where, *y*_*ij*_ is the normalized proportion methylation at probe *i* and sample *j, Sex* is a binary variable (Female=0; Male=1), α_i_ is the proportion methylation in female samples, and *SVs* are significant surrogate variables representing significant variation due to factors other than the covariate of interest (sex). SVs were estimated by the *SVA* R package (version 3.38.0) (30), and 8 significant SVs were found in both discovery and validation datasets.

After fitting Equation 1 to each interrogated cytosine, we computed moderated t-statistics and p-values with the *limma* R package (version 3.46.0) (37), and adjusted the p-values for multiple testing via the Benjamini-Hochberg correction (False Discovery Rate, FDR) (38). Probes with an FDR ≤ 5% were called significantly differentially methylated by sex.

To find differentially methylated regions (DMRs), Eqn. 1 was utilized, and contiguous probes with absolute DNAm differences between sexes of > 0.1 were identified with the *bumphunter* R package (version 1.32.0) (39) (argument: cutoff = 0.1). Statistical significance of the DMRs was assessed by performing linear model bootstrapping with 1000 permutations (by using the *bumphunterEngine* function with the following arguments: nullMethod = ‘bootstrap’ and B = 1000). DMRs with family-wise error rate (FWER) ≤ 0.1 were called significant. To find long-range changes in DNAm (blocks), we utilized Eqn. 1 and the *blockFinder* function of the *minfi* package (33). *blockFinder* collapses neighboring CpGs/probes into a single probe-group measurement, and fits Eqn. 1 for each *i*^*th*^ probe group (thus, *y*_*ij*_ now represents normalized proportion methylation at probe-group *i* and sample *j)*. Similar to DMRs, we required a 0.1 difference in DNAm levels between sexes for DNAm blocks (argument: cutoff=0.1), 1000 linear model bootstrapping permutations were performed (arguments: nullMethod = ‘bootstrap’ and B = 1000), and blocks with FWER ≤ 0.1 were called significant.

DMPs and regions of interest were mapped to genes annotated by GENCODE (version 31). All downloaded 450k datasets were annotated relative to Human Genome version 19 (hg19), so all DNAm analyses were done relative to hg19 for consistency. Whether or not CpGs/probes are associated with promoters or enhancers was determined from Illumina annotation, which is in turn determined using data from the Methylation Consortium (for enhancers) and ENCODE Consortium (for promoters). When determining whether cytosines of interest are proximal to human transcription factors or imprinted genes, a list of human imprinted genes was obtained from geneimprint (www.geneimprint.com), and a comprehensive list of human transcription factors was obtained from a review article by Lambert et al (http://humantfs.ccbr.utoronto.ca/allTFs.php) (31). All analyses were done on R (version 4.0.3).

### Statistical tests

To calculate the significance of differences in number of differentially expressed genes and TFs between datasets, the Chi squared test with continuity correction was utilized. Computations were performed in R (version 4.0.3).

## Data availability

Processed RNA sequencing data (*log*_*2*_*(TPM+1)* values) for all samples that were sequenced for this study is provided in Supplementary Data 1.

## Code availability

All code is deposited on GitHub (https://github.com/nikolayaivanov/PNET_sex_dimorphisms).

## Data Availability

Processed RNA sequencing data (log2(TPM+1) values) for all samples sequenced in this study are provided as Supplementary Data 1.

## Acknowledgements

The authors acknowledge Weill Cornell Scientific Computing Unit Staff for their help with setting up the workflow on the computing cluster.

This work is supported by the National Center for Advancing Translational Sciences of the National Institutes of Health under Award Number TL1TR002386-04 (NAI) and North American Neuroendocrine Tumor Society (NANTS 190177; to IMM). It is also supported by the STARR Consortium (I13-0052), the NIH (R01MH117406, P01CA214274, R01CA249054, R01MH117406), and the Leukemia and Lymphoma Society (LLS) grants (LLS Grant: MCL7001-18, LLS 9238-16, LLS-MCL7001-18) (CEM).

## Author Contributions

Conceived and designed the study: NAI, IMM, CEM. Collected tissue samples: TJF, BMF, IMM. Analyzed data: NAI, KG. Wrote the paper: NAI, IMM, CEM. All authors reviewed, edited, and approved the paper.

