## Supplementary Figures for "Sex-specific transcriptional differences and loss of gene imprinting in pancreatic neuroendocrine tumors"

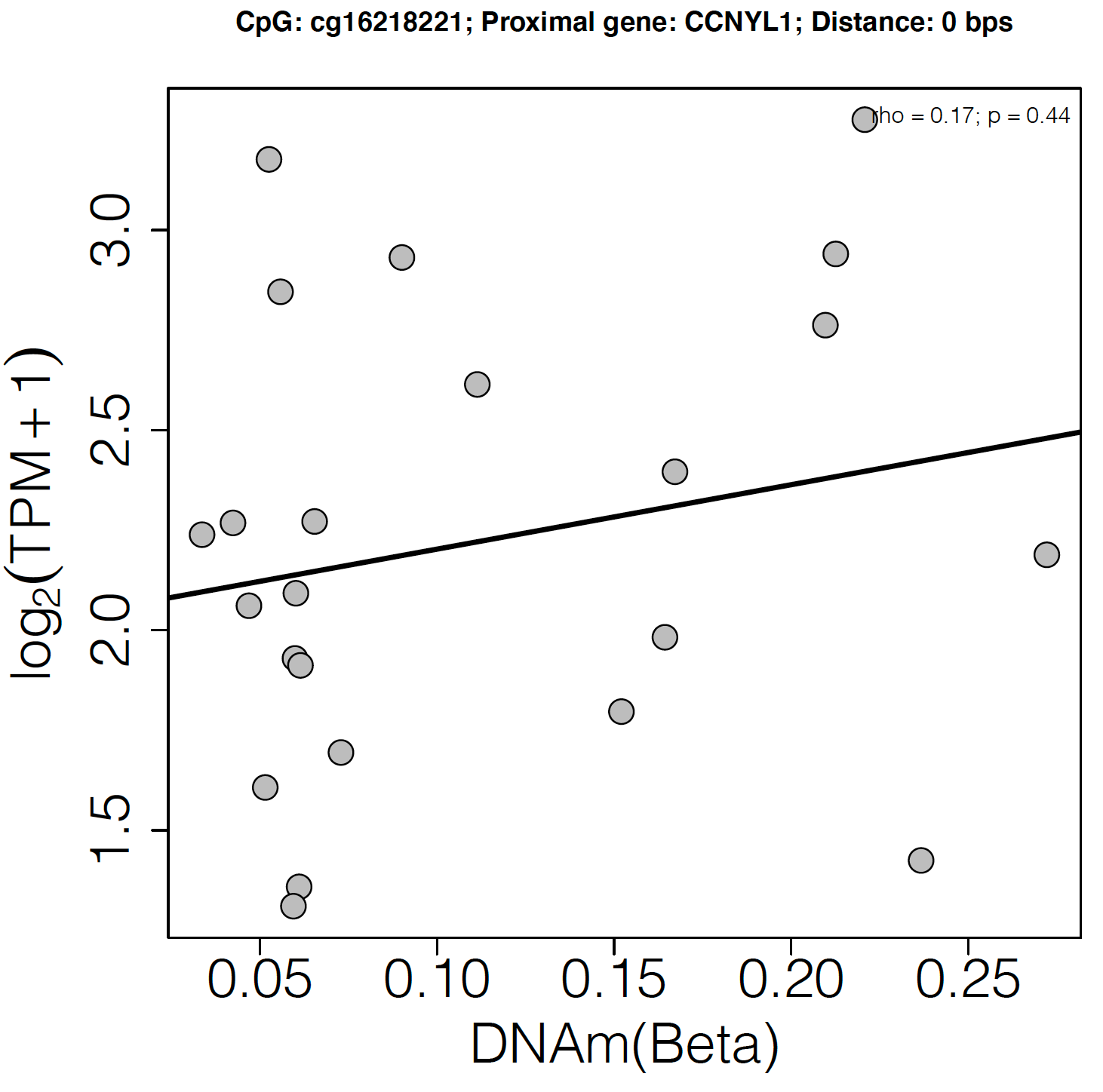

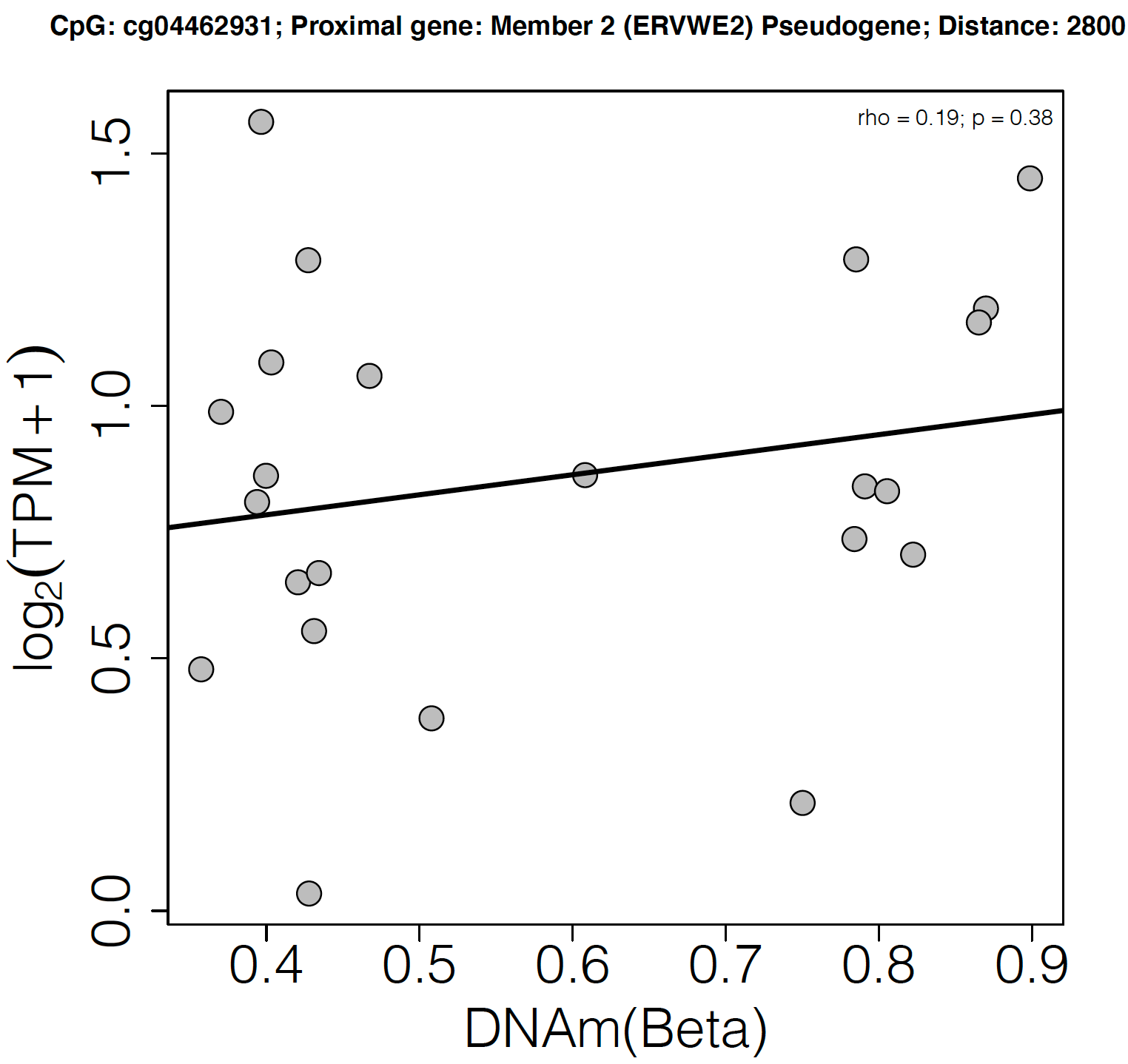

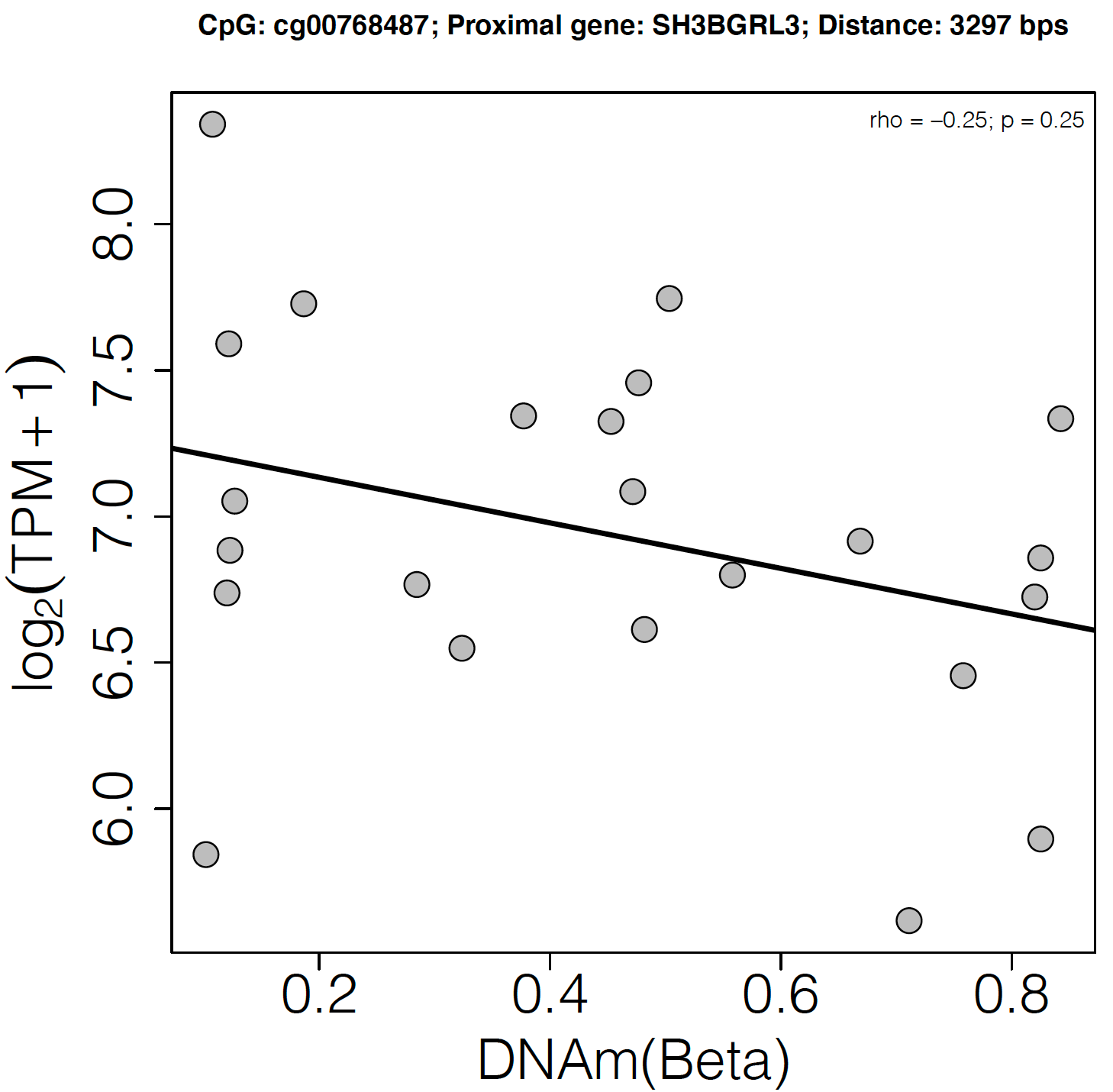

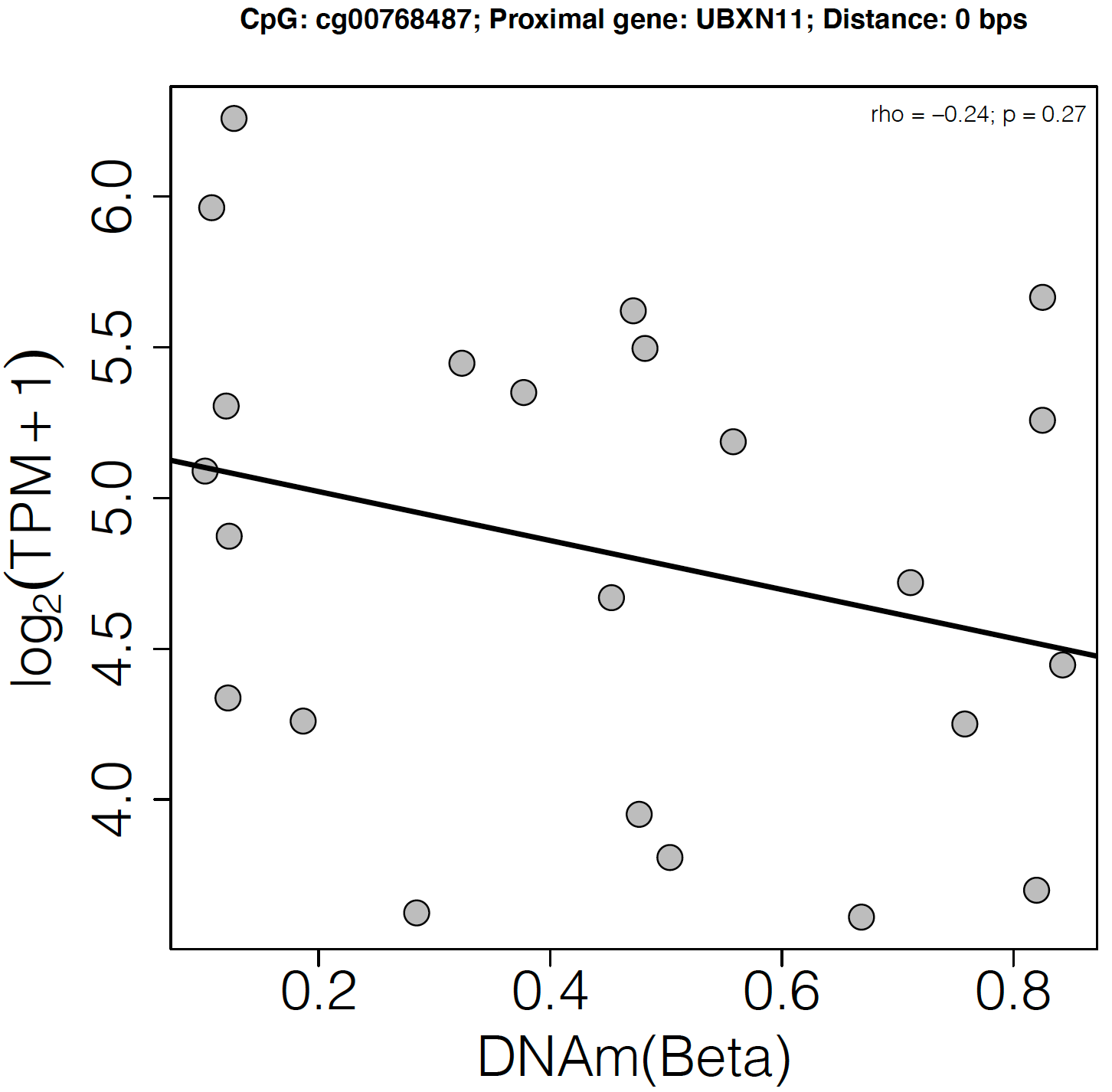

**Supplementary Figure 1.** Correlation between DNA methylation of cytosines differentially methylated by sex in PNETs and genes proximal to them (i.e. overlapping of within 5 kb). Spearman’s rank correlation coefficient (ρ) was used to measure the DNAm – gene expression association.

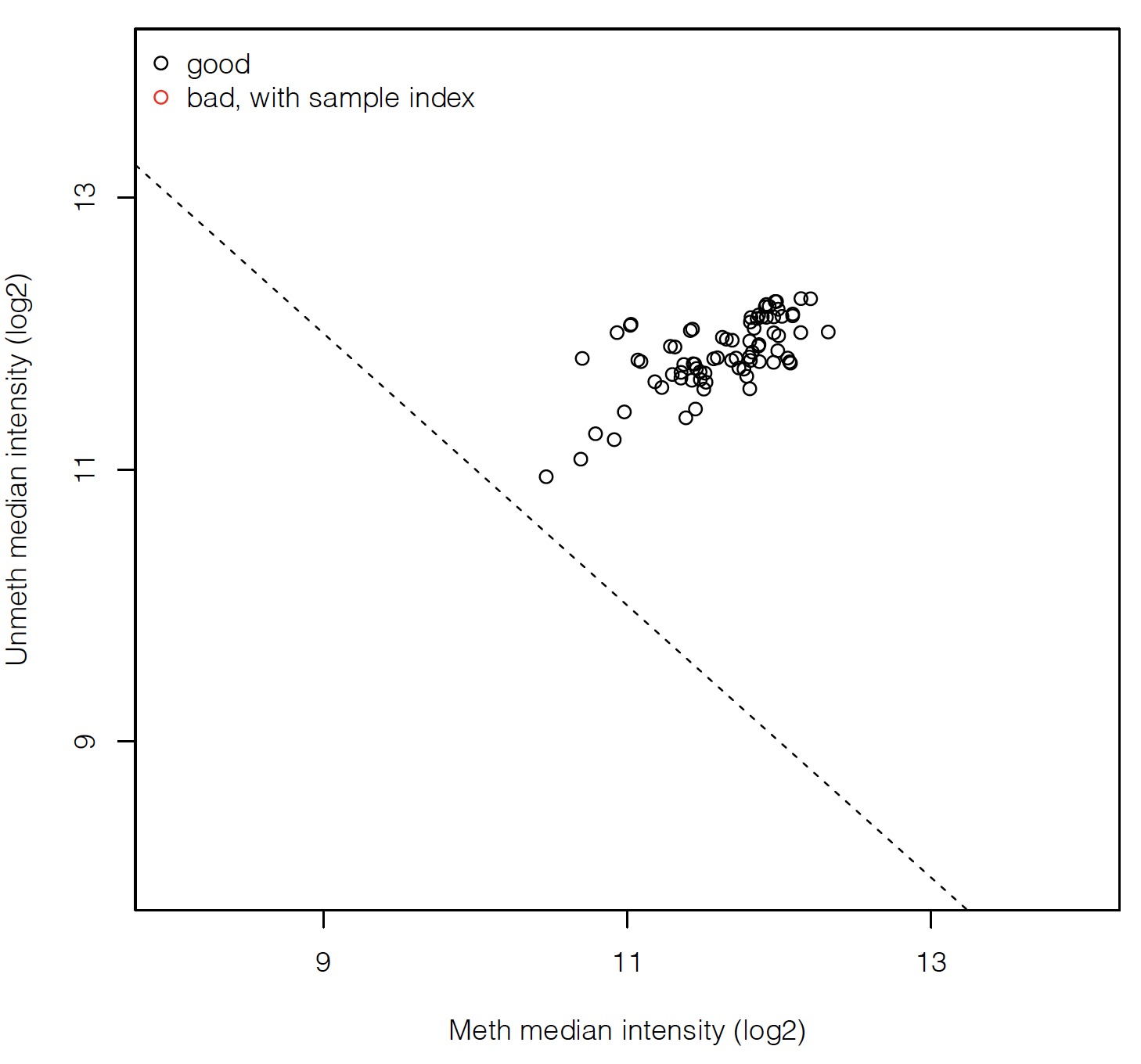

**Supplementary Figure 2**: DNA methylation data quality control. Log_2_ of the median methylated intensity plotted with respect to log_2_ of the median unmethylated intensity. Samples that have low median methylated and unmethylated intensities are typically flagged as likely low-quality samples. None of our samples had to be flagged for low quality.

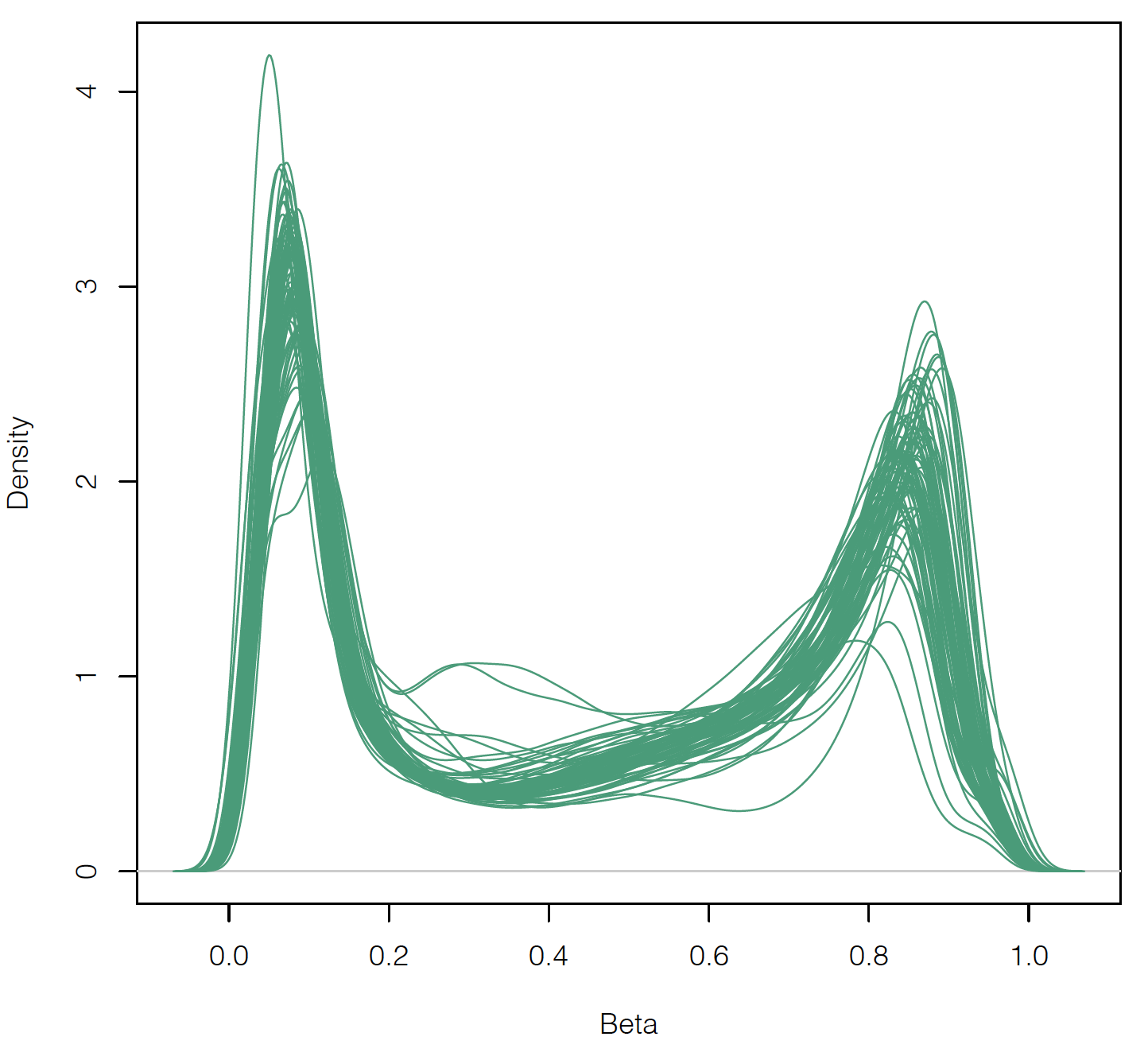

**Supplementary Figure 3**: Beta value densities of all DNA methylation samples. Note that the majority of samples have a bimodal distribution of Beta values, whereby there is enrichment for low Beta values (close to 0), and high Beta values (close to 1), and a relative depletion of intermediate Beta values.
